# Examining Longitudinal Markers of Bladder Cancer Recurrence Through a Semi-Autonomous Machine Learning System for Quantifying Specimen Atypia from Urine Cytology

**DOI:** 10.1101/2023.03.02.23286716

**Authors:** Joshua J. Levy, Natt Chan, Jonathan D. Marotti, Nathalie J. Rodrigues, A. Aziz O. Ismail, Darcy A. Kerr, Edward J. Gutmann, Ryan E. Glass, Caroline P. Dodge, Arief A. Suriawinata, Brock Christensen, Xiaoying Liu, Louis J. Vaickus

**Affiliations:** Emerging Diagnostic and Investigative Technologies, Department of Pathology and Laboratory Medicine, Dartmouth Hitchcock Medical Center, Lebanon, NH, 03766; Department of Dermatology, Dartmouth Hitchcock Medical Center, Lebanon, NH, 03766; Department of Epidemiology, Dartmouth College Geisel School of Medicine, Hanover, NH, 03756; Program in Quantitative Biomedical Sciences, Dartmouth College Geisel School of Medicine, Hanover, NH, 03756; Dartmouth College Geisel School of Medicine, Hanover, NH, 03756; White River Junction VA Medical Center, White River Junction, VT, 05009; UPMC East, Pittsburg, PA, 15146; Cambridge Health Alliance, Cambridge, MA, 02139; Department of Molecular and Systems Biology, Dartmouth College Geisel School of Medicine, Hanover, NH, 03756; Department of Community and Family Medicine, Dartmouth College Geisel School of Medicine, Hanover, NH, 03756

## Abstract

Urine cytology (UC) is generally considered the primary approach for screening for recurrence of bladder cancer. However, it is currently unclear how best to use cytological exams themselves for the assessment and early detection of recurrence, beyond identifying a positive finding which requires more invasive methods to confirm recurrence and decide on therapeutic options. As screening programs are frequent, and can be burdensome, finding quantitative means to reduce this burden for patients, cytopathologists and urologists is an important endeavor and can improve both the efficiency and reliability of findings. Additionally, identifying ways to risk-stratify patients is crucial for improving quality of life while reducing the risk of future recurrence or progression of the cancer. In this study, we leveraged a computational machine learning tool, AutoParis-X, to extract imaging features from UC exams longitudinally to study the predictive potential of urine cytology for assessing recurrence risk. This study examined how the significance of imaging predictors changes over time before and after surgery to determine which predictors and time periods are most relevant for assessing recurrence risk. Results indicate that imaging predictors extracted using AutoParis-X can predict recurrence as well or better than traditional cytological / histological assessments alone and that the predictiveness of these features is variable across time, with key differences in overall specimen atypia identified immediately before tumor recurrence. Further research will clarify how computational methods can be effectively utilized in high volume screening programs to improve recurrence detection and complement traditional modes of assessment.

## Introduction

Urothelial carcinoma ranks ninth worldwide in cancer incidence as the seventh most common malignancy in men and seventeenth in women ^1–3^. In the United States, urinary bladder cancer (UBC) is the fourth most common cancer in men and tenth in women. Of urothelial cancer cases, most are forms of UBC at approximately 90%, while upper tract urothelial carcinomas account for 5-10% of malignancies ^4–7^. The 5-year relative survival rates for UBC patients range from 97% at Stage I to 22% at Stage IV ^8–11^. Most UBC incidences (75-85%) are non-muscle invasive (NMIBC) at first diagnosis, of which 70% register as pTa (noninvasive papillary carcinoma), 20% as pT1, and 10% as carcinoma in situ (CIS) lesions, pTis. The prognosis of NMIBC is generally favorable, although 30-80% of cases will recur and 1-45% of cases will progress to muscle invasion within five years ^12^. As a result, NMIBC is treated as a chronic disease with a variety of oncological outcomes that require frequent follow-ups for monitoring and repeated treatments, giving it the highest cost-per-patient from diagnosis to death of all cancers ^13^.

The standard approach to patients with symptoms suggestive of UBC involve a combination of urine cytology, cystoscopy (potentially with tissue biopsy(s)), and immunocytochemical and molecular studies with longitudinal follow-up for negative and atypical findings ^14–23^. After a positive diagnosis of UBC, urine cytology remains an essential longitudinal monitoring tool for patients. However, urine cytology suffers from susceptibility to issues such as specimen quality, inter/intra-observer variability, and ‘hedging’ towards atypical diagnosis, making it a semi-qualitative assessment and vulnerable to individual biases ^24–28^. Such factors restrict the predictive value of urine cytology therefore increasing reliance on invasive cystoscopy.

Cytology specimens have historically been tedious to screen, in part due to the sheer volume of specimens to examine, resulting from regular periodic follow-up and the highly variable specimen cellularity. While positive and negative urine cytology specimens are easier to classify, atypical and suspicious urine samples are more challenging and feature poor inter-observer reproducibility. In recent years, The Paris System for Reporting Urinary Cytology (TPS), published in 2016 and updated in 2022, has established itself as the widely accepted classification system for UBC screening ^24,29,30^. It devised to tackle the challenges posed by atypical urines and improve reproducibility ^31,32^. Computer algorithms such as the AutoParis system were designed to ameliorate many of these screening challenges/burdens to make urine cytology quantitative by employing machine learning techniques that can mimic rapid examination with TPS criteria ^33–39^. AutoParis, and its latest iteration, AutoParis-X, calculate an Atypia Burden Score (ABS) after cross-tabulating several cellular and cluster-level subjective and objective indicators of atypia ^34,40^.

As bladder cancer recurrence is a significant concern for patients and healthcare providers, various methods have been developed to predict and monitor the likelihood of recurrence. While computer-aided assessment of the primary tumor has been shown to be predictive of likelihood for recurrence ^41,42^, this examination presents only a snapshot in time, which could be augmented by repeated urine cytology exams ^43–45^. However, there is currently little to no research on how repeat urine cytology exams can be leveraged to derive longitudinal markers of recurrence ^46–49^. Assessing the prognostic capacity of imaging predictors in urine specimens for the treatment of bladder cancer can have great benefits in reducing clinician workload, improving reproducibility, reducing human error, and lowering treatment cost, in part because cytology predictors can serve as an “early warning system” for which patients require the most attention/care ^50^. In this specific work, we investigate the potential of using machine learning from urine cytology in predicting recurrence among a cohort of patients ^40^.

## Methods

### Methods Overview

In this section, we summarize the approaches taken to assess the ability to predict time to recurrence from image-derived UC predictors:

1. Retrospective review identifies cases with varying follow-up and number of recurrences.
2. Slide images are scanned (**Figure 1A**) and imaging predictors are extracted from each whole-slide image (WSI) (**Figure 1B**) using AutoParis-X, which improved upon techniques introduced by AutoParis ^34,40^.
3. *Fixed predictors* are constructed by aggregating quantitative cytological exam information across distinct collection periods (i.e., collection time; **Figure 1C**); Cox proportional hazards models are developed to predict recurrence risk and compare with manual assessments (UC Class) and tumor grade/stage/type (histology) ^51,52^.
4. *Dynamic predictors* are constructed by utilizing the imaging predictors of each individual cytology exam; these predictors vary with time (time-varying covariates) and their effects are reported across different time periods through time-varying coefficient Cox models (**Figure 1D**) ^53,54^.
5. Models are interpreted by regression coefficients (i.e., hazard ratios), concordance statistics, and clustering time series, which shows how imaging predictors vary across time for low-risk / high-risk patients with commensurate statistical modeling (e.g., hierarchical beta regression) ^55,56^.

**Figure 1:**
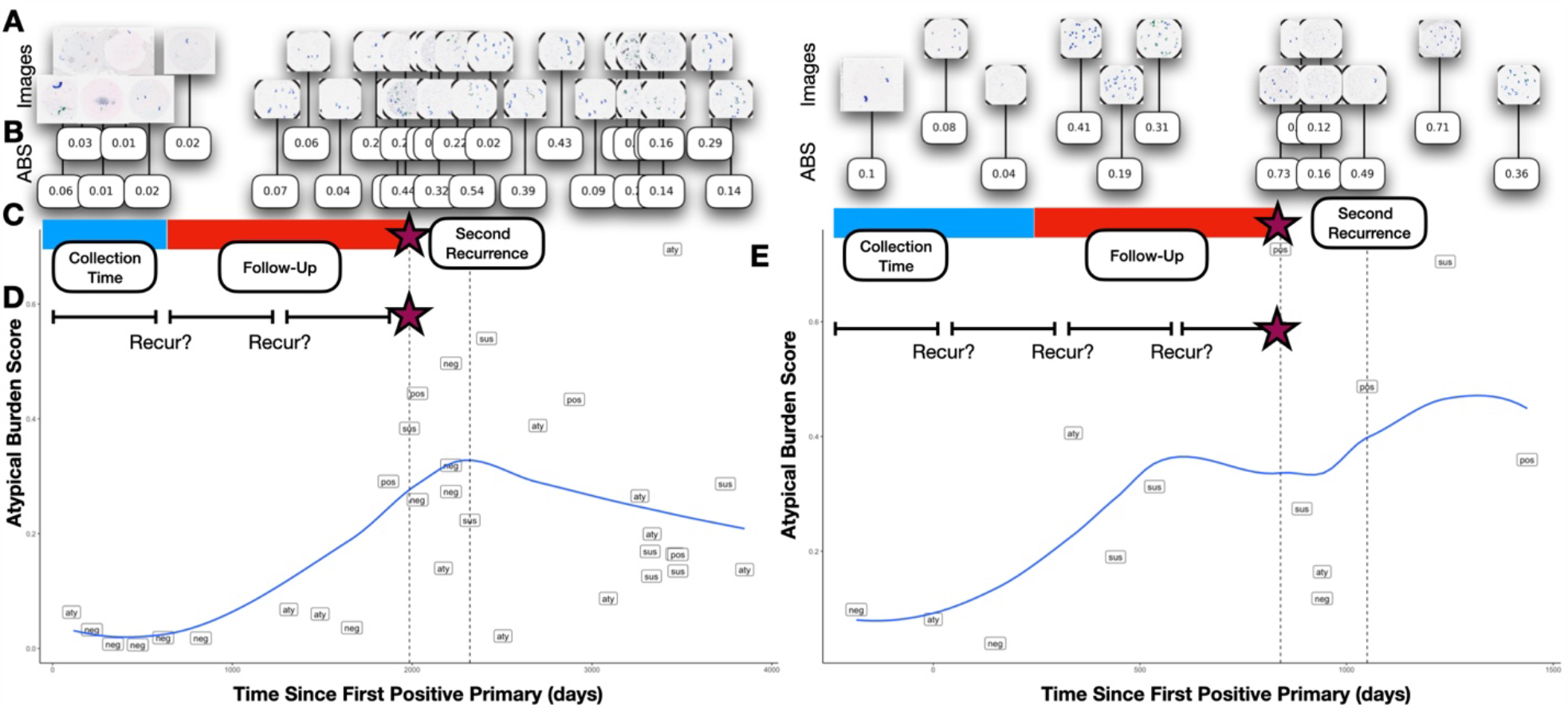
Study Overview: **A)** UC images are acquired for patients across the study period and are processed using **B)** AutoParis-X, which extracts imaging predictors, e.g., ABS; **C)** Imaging predictors are aggregated across collection periods to form *fixed predictors* which are then used to assess time-to-recurrence using Cox models; **D)** Imaging predictors were also studied *dynamically* considering results/extracted features from individual tests and their recurrence potential; risk of recurrence was also studied within specific time periods to demonstrate how the importance of these predictors varies with time; **E)** Scatterplots for two patients with time from the first positive primary versus the Atypia Burden Score as assessed using AutoParis-X; points were labeled by the UC categories assigned through manual examination of urine cytology

### Specimen Collection

A total of 1,259 urine specimens collected from 135 bladder cancer patients at Dartmouth-Hitchcock Medical Center between 2008 and 2019 were retrieved, after institutional review board approval. The median number of specimens per patient was 8, with an interquartile range of [8-13] (**Figure 1A**). Several patients were omitted due to insufficient follow-up or significant left-censoring which precluded assessment. The specimens were prepared using ThinPrep® (Hologic, Marlborough MA) and Papanicolaou staining before being examined microscopically ^57^. They were then scanned with a Leica Aperio-AT2 scanner at 40x resolution, resulting in full-resolution SVS files (70% quality JPEG compression) representing whole slide images. The slides were manually focused on a single plane during scanning, without the use of z-stacking ^58^. Patient and slide-level characteristics from the retrospective cohort are provided in **Table 1**. All slides were evaluated by five cytopathologists to provide diagnoses based on The Paris System criteria (negative, atypical, suspicious, positive). Separately, patient characteristics, e.g., hematuria, prior treatments (e.g., BCG– Bacillus Calmette-Guerin or mitomycin) were recorded in a secure database ^59^. Time to recurrence was determined as indexed from the date of the first positive primary tumor as determined through histological examination. Individuals were right censored based on last known histological follow-up ^60^.

**Table 1:**
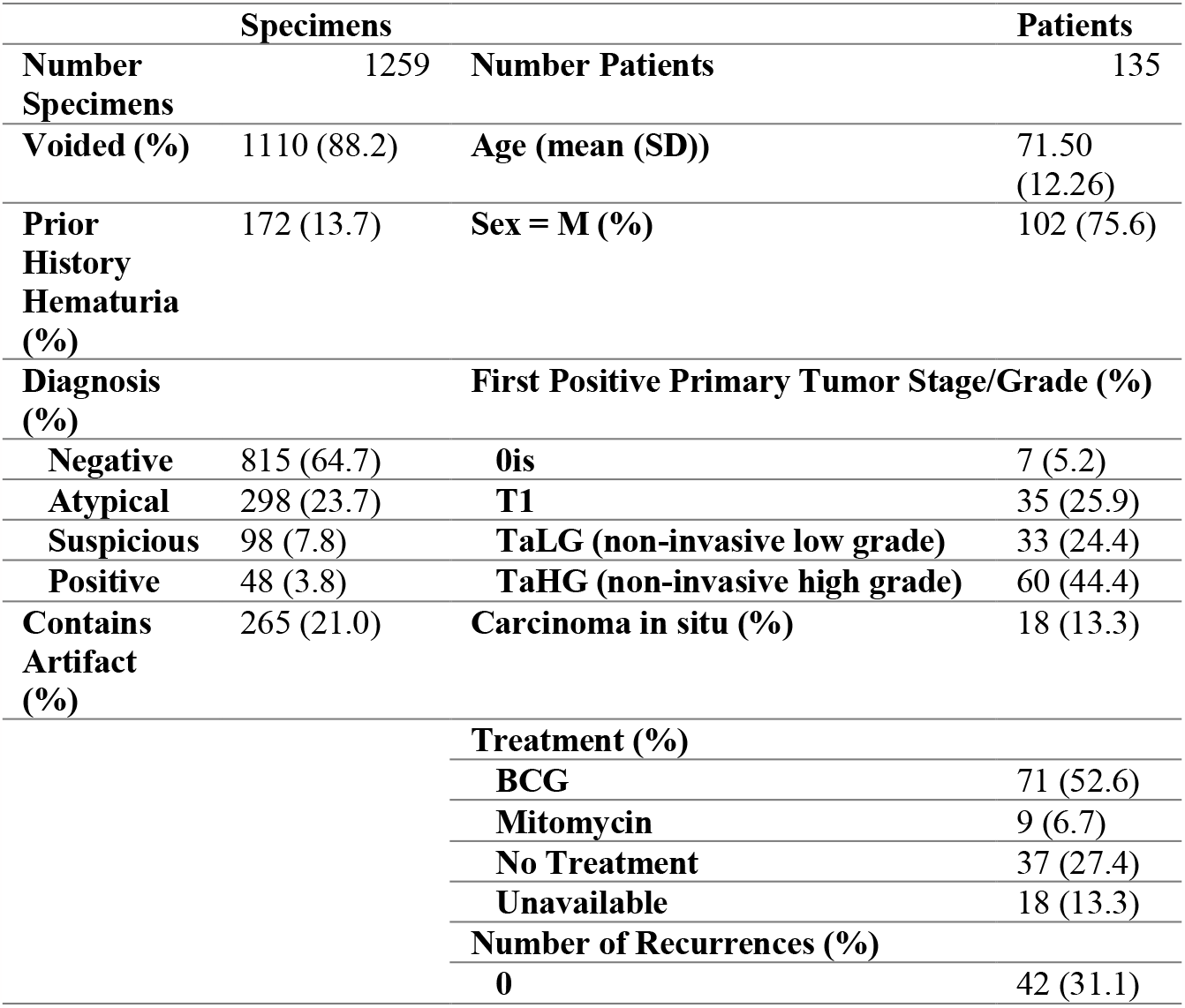
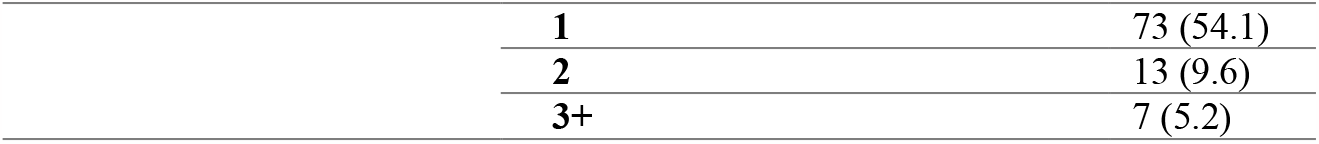
Patient and specimen characteristics

| Specimens |  | Patients |  |
| --- | --- | --- | --- |
| **Number Specimens** | 1259 | **Number Patients** | 135 |
| **Voided (%)** | 1110 (88.2) | **Age (mean (SD))** | 71.50 (12.26) |
| **Prior History Hematuria (%)** | 172 (13.7) | **Sex = M (%)** | 102 (75.6) |
| **Diagnosis (%)** |  | **First Positive Primary Tumor Stage/Grade (%)** |  |
| **Negative** | 815 (64.7) | **0is** | 7 (5.2) |
| **Atypical** | 298 (23.7) | **T1** | 35 (25.9) |
| **Suspicious** | 98 (7.8) | **TaLG (non-invasive low grade)** | 33 (24.4) |
| **Positive** | 48 (3.8) | **TaHG (non-invasive high grade)** | 60 (44.4) |
| **Contains Artifact (%)** | 265 (21.0) | **Carcinoma in situ (%)** | 18 (13.3) |
|  |  | **Treatment (%)** |  |
|  |  | **BCG** | 71 (52.6) |
|  |  | **Mitomycin** | 9 (6.7) |
|  |  | **No Treatment** | 37 (27.4) |
|  |  | **Unavailable** | 18 (13.3) |
|  |  | **Number of Recurrences (%)** |  |
|  |  | **0** | 42 (31.1) |

|  |  |
| --- | --- |
| **1** | 73 (54.1) |
| **2** | 13 (9.6) |
| **3+** | 7 (5.2) |

### Using AutoParis-X to Derive Imaging Predictors of Recurrence

AutoParis-X is a tool for automated assessment of cytology specimens that was developed using the Python programming language and the PyTorch and Detectron2 frameworks, with statistical and machine learning models implemented in Python and R ^40,61–64^. In brief, this tool:

1. Utilizes connected components analysis to isolate individual cells and cell clusters
2. A neural network-based cell border detection model called BorderDet isolates urothelial cells within clusters and identifies dense overlapping cell architectures ^65^.
3. Additional morphometric measures are derived for cell-type classification and atypia estimation ^34,40^.
4. A convolutional neural network called UroNet filters out any objects which are not urothelial cells ^34,40^.
5. A segmentation neural network method called UroSeg estimates the nuclear-to-cytoplasm ratio ^34,40^.
6. A convolutional neural network called AtyNet scores cells for subjective markers of atypia ^40^.
7. A machine learning classifier estimates the Atypia Burden Score (ABS) which integrates cell and cluster-level scores and other demographic and specimen characteristics into a summary measure of overall specimen atypia ^66–68^.
8. In addition, hierarchical regression models identified important indicators of atypia, and graphical displays were generated through an interactive web application utilized by our team of cytopathologists ^69,70^.

A description of slide level measures and ABS scores, listed in **Supplementary Table 1**, which were derived for each specimen in this cohort.

### Recurrence Prediction

Time to recurrence was predicted using both traditional cytological measures and AutoParis-X derived imaging features (**Figure 1B**), controlling for age and sex, prior treatment, tumor grade, medical history, etc., where possible– e.g., treatment information was largely excluded from multivariable modeling due to missingness and uncertainty in treatment time.

#### Fixed recurrence predictors

First, we aggregated imaging/cytology statistics (e.g., average number of atypical cells) for cytology exams before/at the primary diagnosis date or within a specific time frame after the primary diagnosis date (i.e., *collection time*) (**Figure 1C**). It is important to ensure that data is collected up to a specific date in order to accurately assess risk for new patients. This is because collecting data beyond this point would introduce information about the future and potentially bias the results. To ensure that the findings remain applicable, data for new patients must be collected only up to the defined collection time. Cases were excluded if events/censoring occurred before this collection window and recurrence times were adjusted as appropriate (i.e., delayed entry) to avoid endogeneity. We denote predictors during this time period as *fixed predictors*. Fixed predictors were modeled using multivariable cox proportional hazards models ^71^:

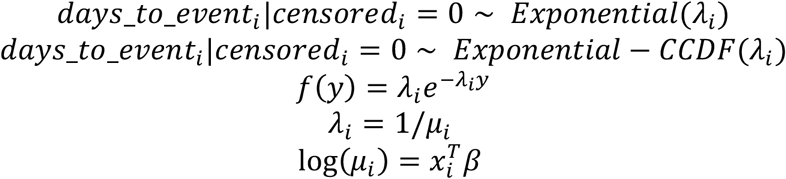

The predictive performance of leveraging fixed (i.e., collected) UC imaging predictors was compared to the histological examination, e.g., tumor grade/stage and whether the tumor was carcinoma in situ (Cis). Separate cox models were fit to the imaging predictors alone, tumor grade and carcinoma in situ, and both, adjusting for age and sex. Models were compared through partial likelihood ratio testing, which would indicate whether imaging predictors alone were more informative than the histological findings (**H**_**1**_**: Imaging>Grade+Cis**) and separately whether the imaging predictors supplemented tumor grade information to add additional predictive capacity (**H**_**1**_**: Imaging+Grade+Cis>Grade+Cis**). We separately reported the hazard ratios for the imaging predictors after adjusting for tumor grade/stage and Cis. Results were compared at all collection times. We did not adjust models for whether the patients had chemotherapy due to unreliability in recording patient start date and adherence, though this information was recorded in the demographic tables for additional context.

#### Dynamic recurrence predictors

Time-dependent predictors (denoted as *dynamic predictors*) were modeled using cox proportional hazards models which allowed repeat measures by patient. These predictors were modeled with and without time varying effects (similar to estimating multiple survival curves across discrete time intervals) (**Figure 1D**), which reports changes to the relationship between predictors and recurrence as a function of time (i.e., certain intervals may be more predictive of recurrence) ^53^.

Individual predictors were modeled in a univariable setting, adjusting only for age and gender. These variables were combined into multivariable models. Predictor selection was accomplished using the variance inflation factor (VIF) after fitting the survival models and iteratively removing predictors until the largest VIF score was less than 6.5 ^72^. We had also performed LASSO predictor selection but opted for VIF as these models outperformed LASSO ^73^. Concordance statistics (C-index; as reported using the *survival* R package) were reported for the univariable and multivariable models, along with hazards ratios, confidence intervals and p-values. For the time-varying effects, hazards ratios and their statistical significance were reported across time for individual predictors and overall across many predictors ^54^. Hazard predictions were dichotomized into low and high risk and *fixed predictors* were visualized using Kaplan Meier plots using the *survminer* package (R v4.1) ^74^.

### Studying Trajectories of ABS Scores

After fitting the cox models, we additionally sought to uncover longitudinal patterns of atypia related to high recurrence risk (**Figure 1E**). This was accomplished by clustering the trajectories of ABS scores across time using dynamic time warping (DTW). DTW was used to construct a distance matrix between individual patient trajectories, which were reduced into two features per patient using multi-dimensional scaling using the *scikit-time* library (Python v3.8) and *reticulate* package (R v4.1) ^75,76^. Separately, the patients were clustered using hierarchical clustering of the DTW distance matrix via the *hclust* function (R v4.1). Cases were omitted if they did not contain at least two points. Associations between the DTW clusters and features were identified through generalized linear mixed effects modeling. The average ABS score was visualized across time, aggregated for low-risk / high-risk patients and separately for the derived clusters at binned time periods. Beta hierarchical regression models with post-hoc comparison via emmeans were used to report how ABS differed between high and low risk patients across time ^55,56,77^:

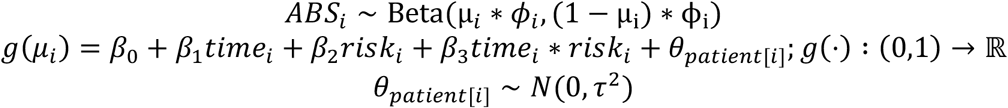

We identified several patients who had multiple recurrences. We visualized changes in ABS before and after recurrences by creating scatter plots of ABS versus time. We fit a hierarchical beta regression model to depict overall changes in ABS score across time between patients’ first and second recurrences, with similar hierarchical beta regression models fit, excluding *risk* from the model.

## Results

### Recurrence Predicted from Fixed Predictors

Fitting cox proportional hazards models at various collection times, we found there was moderate ability to predict recurrence using UC imaging predictors (**Figure 2C,D; Supplementary Figure 1**; **Supplementary Table 2**). When only collecting cytological information up to the first positive primary (collection time = 0 days), imaging and manually assessed UC class predictors yielded a C-index of 0.672. Overall, imaging predictors were more informative than manual cytological examination (**Supplementary Table 2**, see “% Outperform UC Class”, number of imaging predictor with better performance than manual examination). The predictiveness of the UC imaging predictors increased when predictors were aggregated across larger time intervals / collection times, for instance yielding a C-index of 0.77 when collecting quantitative cytological information over the first 180 days after the first positive primary (collection time = 180 days). Collecting cytological information past this point in time and aggregating yielded marginal to no additional information on recurrence. The imaging variables differed significantly in their predictive capacity. Surprisingly, imaging features extracted from urothelial cell clusters proved remarkably predictive (C-index for: number of atypical cell clusters = 0.733; number of dense cell clusters = 0.748 at collection time 180 days) as opposed to variables which correlate more closely with UC Class (e.g., ABS).

**Figure 2:**
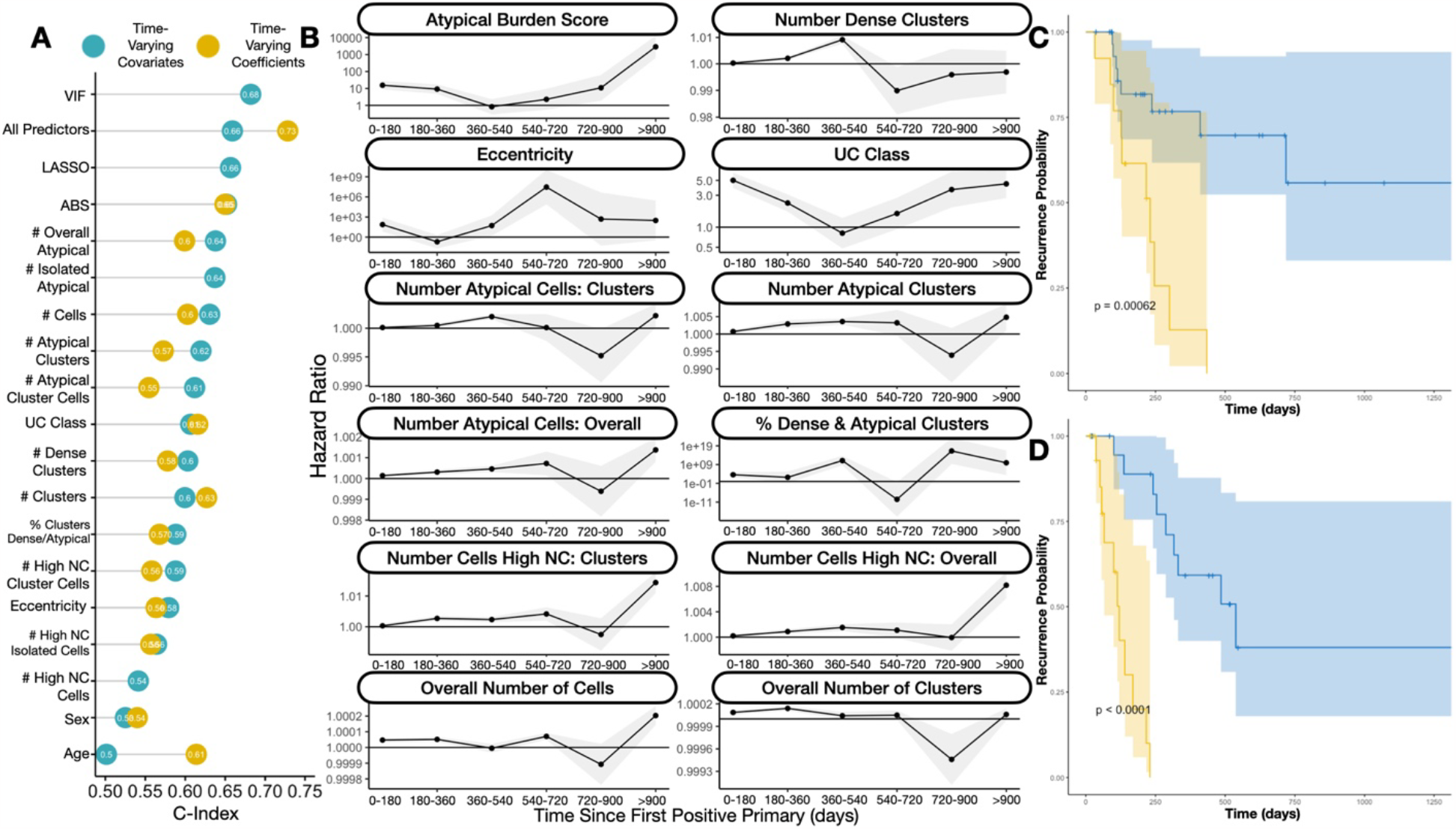
Findings from Recurrence Risk Models: **A)** Dot chart indicating concordance statistics for each of the imaging predictors for the *time-varying covariate* and *time-varying effects* cox proportional hazards models; UC class stands for category assigned via manual examination by the cytopathologist; VIF and LASSO refer to multivariable models with the respective predictor selection methods; All/Overall predictors refers to multivariable models with all imaging predictors; **B)** Ribbon plot illustrating hazard ratios and confidence intervals for univariable *time-varying effects* cox proportional hazards model for individual imaging predictors, demonstrating differing associations with recurrence at distinct time intervals; **C)** Kaplan-Meier plot and rank-based statistic for *fixed imaging predictors* collected before or up to the date of the first positive primary, reported for low (blue) and high (yellow) risk patients as assessed using the Cox model; **D)** similar KM plot for patients with 90 days of follow-up information collected, predicting recurrence risk after this collection period

Imaging predictors extracted from cytology and separately in conjunction with risk assessment models based on tumor grade/type were more informative for recurrence risk prediction than that derived from tumor grade/type alone (**Supplementary Figure 2; Supplementary Table 3**), as assessed through partial likelihood ratio testing ^78,79^. At nearly every collection interval, imaging predictors demonstrated statistically significantly better predictive capacity than tumor grade/type alone and effects from the imaging predictors were highly statistically significant, even after adjusting for tumor grade/type (**Supplementary Table 3**).

### Recurrence Predicted from Dynamic Predictors

When considering all individual cytology exams dynamically over time (*time-varying covariates*) and not aggregating across distinct time windows, imaging predictors corresponded with recurrence with a C-index of 0.66 (**Figure 2A; Supplementary Table 2**). The Atypical Score (C-index=0.65) was more predictive than UC Class (C-index=0.58) using this approach. Fitting recurrence models, allowing effects of different predictors to vary– these time varying effects were reported for each distinct time period (*time-varying effects*; association between variables and recurrence risk updated every half year; **Figure 2B**), achieved an overall C-index of 0.73, greater than that offered by the *time-varying covariates*. The Atypical Score (C-index=0.65) was still more predictive than UC Class (C-index=0.62) using this approach (**Supplementary Table 4**). The association between individual imaging predictors and recurrence risk varied across these intervals (**Figure 2B; Supplementary Table 5**). For instance, ABS and UC Class were highly positively associated with recurrence risk during the first year and after the second year of follow-up (**Figure 2B; Supplementary Table 5**). As another example, the number of atypical cells and atypical clusters demonstrated their greatest association with recurrence risk at intermediate intervals (e.g., 180-540 days) (**Figure 2B; Supplementary Table 5**).

### Trajectory Cluster Analysis

We sought to study the trajectories of specimen atypia from the first positive primary to the first recurrence. Time series clustering yielded three independent clusters (**Figure 3A**). The red cluster (**Figure 3A**) revealed the tendency of patients to exhibit a decrease in specimen atypia immediately after the positive primary (likely resulting from previous treatment), followed by a sharp increase in specimen atypia thereafter. Patients deemed high risk by the Cox models (**Figure 3B**,**C**) initially have a low atypical burden, similar to the low risk group. However, over time after the positive primary, the discrepancies in specimen atypia increase substantially (**Figure 3B; Supplementary Table 6**). When counting down backwards from the date of first recurrence, we see that specimen atypia increases steadily from both low and high risk patients prior to the first recurrence. Within 3-4 months prior to the first recurrence, specimen atypia for the low-risk patients decreases while continuing to increase for the high risk patients (**Figure 3C; Supplementary Table 6**).

**Figure 3:**
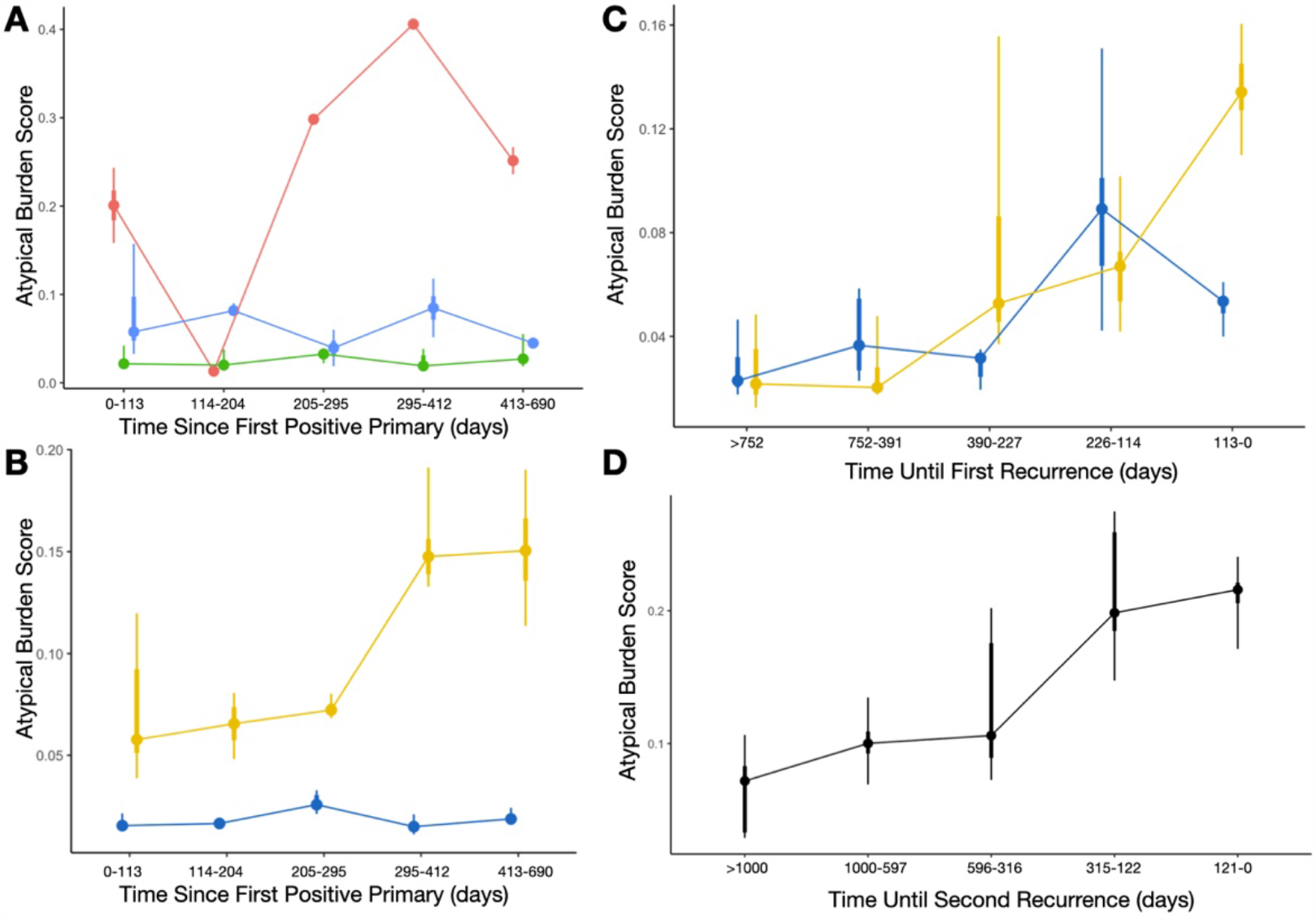
Atypia Burden Scores reported across time, aggregated across distinct time periods using point interval plots: **A)** Each curve/color represents ABS scores from patients belonging to three different temporal trajectories (red, blue, green clusters), determined using the time series clustering and summarized using the aggregate statistics for each time period; **B)** Each curve is colored based on low (blue) and high (yellow) risk patients, measured from the time since first positive primary; **C)** Comparing ABS scores between low/high risk patients, similar to the previous plot, with cytological exams grouped by days until the first recurrence instead of from the date of the first positive primary; **D)** ABS scores, combined across distinct time periods, for patients from the first until the second recurrence, grouped by the days until the second recurrence, demonstrating increasing atypia prior to the recurrence finding

These trends were similarly identified for patients who had a first recurrence who would go onto have a second recurrence (**Figure 3D; Supplementary Table 6; Supplementary Figure 3**). A statistically significant increase in overall specimen atypia over time was identified during this interval between the first and second recurrences (**Supplementary Table 6**). The Atypia Burden Scores plotted across time from positive primary date for patients with multiple recurrences can be found in **Supplementary Figure 4**, though an in-depth assessment is outside of the scope of this study.

## Discussion

Bladder cancer has a high rate of recurrence, which requires frequent follow up screening and monitoring. By using advanced computer algorithms, it is possible to create a non-invasive, semi-autonomous system that can analyze repeat cytology exams and provide highly precise markers of specimen atypia ^34,36,40^. This approach can improve our understanding of how bladder cancer progresses and recurs, as well as identify patterns that indicate early detection of recurrence. This study sought to investigate the potential utility of such an approach, made possible by the AutoParis-X tool, which can facilitate rapid examination of cytology specimens ^40^. Imaging predictors derived using AutoParis-X such as the Atypia Burden Score and other sub-scores (e.g., number of atypical clusters) were followed across time for patients and were aggregated across distinct time periods and studied *dynamically* to predict bladder cancer recurrence.

The principal findings from our study are twofold: 1) urine cytology exam results can inform recurrence risk, and imaging predictors extracted through the use of machine learning can be more informative of recurrence than manual cytological and/or histological examination alone; and 2) the predictive value of imaging predictors extracted from UC exams varies across time (both in terms of combining information from previous exams and real-time predictiveness of time-variant predictors). Our findings support and add to previous studies showing that preoperative urine cytology examination can predict recurrence ^80–82^. We also found that collecting and combining cytological information with summary statistics within the first six months after the positive primary diagnosis is important for assessing recurrence risk for patients who have not yet recurred. While there are several other machine learning techniques which have been developed to perform histological assessments of recurrence risk from the primary site at the time of resection, cytological assessments are far less invasive (requiring the patient to simply void into a collection cup in most cases) ^41,83^. Due to routine screening via UC, more information is available, which when assessed in totality, can be highly predictive of recurrence.

It is important to consider how information from cytological and histological examinations can be used together to provide more comprehensive assessment of risk. The use of imaging predictors extracted from cytology, both alone and in combination with tumor grade/type, provided more useful information for predicting recurrence risk compared to relying on tumor grade/type alone, as determined through partial likelihood ratio testing. The combination of cytological and histological assessments is especially pertinent for patients who have undergone a tumor resection and are identified to be at high risk from both cytology and histology.

There are limitations to this study. For instance, there is still ample room to improve the AutoParis-X algorithm, which can impact the reliability of these predictors ^40^. Furthermore, we have not studied its utility in augmenting medical diagnostic decision-making in conjunction with the cytopathologist ^84–88^. Changes in specimen preparation across the past decade and a half may have impacted imaging predictors estimated using AutoParis-X. We used the last histological follow up exam with a negative finding as a right censoring event for patients in this cohort who did not ultimately develop recurrence ^89^. As this was a retrospective cohort study with sporadic follow-up (typically every three months as specified by guidelines), it was challenging to identify suitable follow-up and censorship criteria. Furthermore, death may present a competing risk to recurrence, which could potentially bias effect estimates. While methods do exist to account for competing risks, relevant statistical methods and their computational implementations are underdeveloped and inaccessible in the context of time-varying covariates and effects^90,91^. These limitations will be improved upon in further assessments of this tool and these study findings should be interpreted in the context of an exploratory analysis. The study cohort was restricted to individuals from Northern New England and findings are applicable to this population– expansion of this study to large, diverse study cohorts from geographically disparate regions will improve the generalizability of these findings.

In the future, we plan to leverage additional machine learning techniques which are suitable for recurrence prediction. For instance, tree-boosting approaches and deep learning models exist which are well-suited for the study of longitudinal / time-to-event data ^92–105^. They can reveal interactions between predictors for use in statistical modeling as well as identify cytology exams / timepoints which are most informative of recurrence ^106^. These are estimated dynamically using sophisticated computational heuristics and are an area of future follow-up.

The results of this study highlight the need for further research comparing the performance of the AutoParis-X system with other non-invasive methods for assessing the potential for bladder cancer recurrence. Many promising approaches make use of various molecular assays developed for liquid biopsies, and several screening programs have also been developed that use a combination of different assays to assess the potential for recurrence ^107–114^. These should be considered for comparison when attempting to roll out potential screening systems/guidelines. While early detection of recurrence is important, it is currently unclear what the next steps should be in terms of treatment and management given the adoption of computational systems for real-time recurrence assessment ^35,39,115–117^. This is an area that requires further research.

Furthermore, there are a wide-range of epidemiological studies which could benefit from incorporating cytological information. For instance, exposure to high levels of arsenic in drinking water and cigarette smoking are associated with bladder cancer risk and could benefit from being studied in conjunction with advanced computational methods for urine cytology ^118–123^.

## Data Availability

All data produced in the present study are available upon reasonable request to the authors.

## Conclusion

This study sought to investigate the potential benefit of using computer algorithms to extract highly quantitative, longitudinal cytological features can be used to inform the risk of recurrence for bladder cancer patients. We found that image predictors extracted using the AutoParis-X system were indeed associated with tumor recurrence, in many cases more so than traditional modes of cytological/histological examination, and that the importance/predictiveness of these predictors varied across time from the positive primary. While this study demonstrates the potential utility for computerized systems to supplement and make use of screening programs with a large number of follow up visits, further research is warranted to better understand how these systems can be integrated into such screening programs.

## Appendix

**Supplementary Figure 1:**
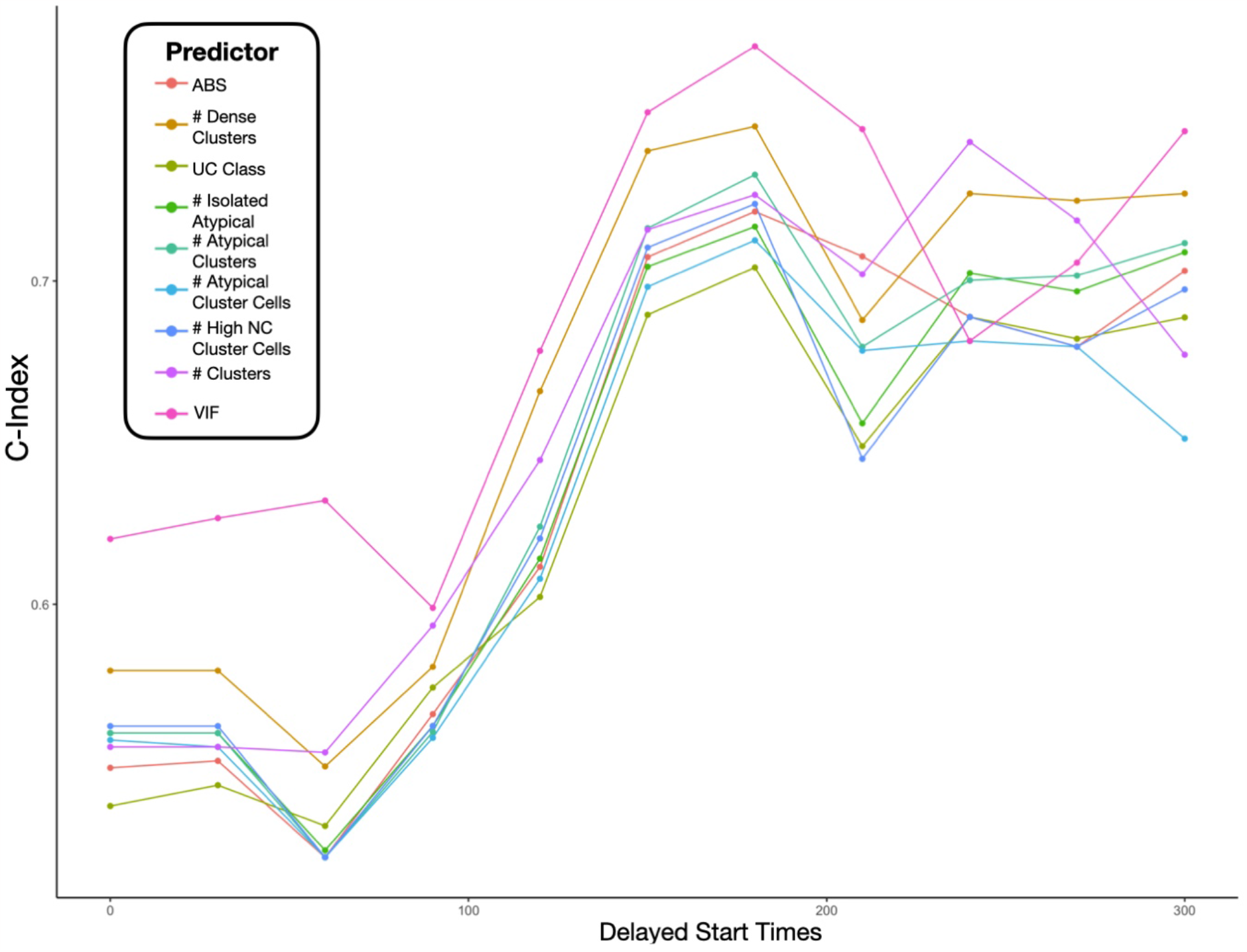
**C-index for specific imaging / manual cytology exam results**, reported based on different collection periods/times (days) prior to the recurrence risk follow-up period; only select AutoParis-X measurements of interest were reported

**Supplementary Figure 2:**
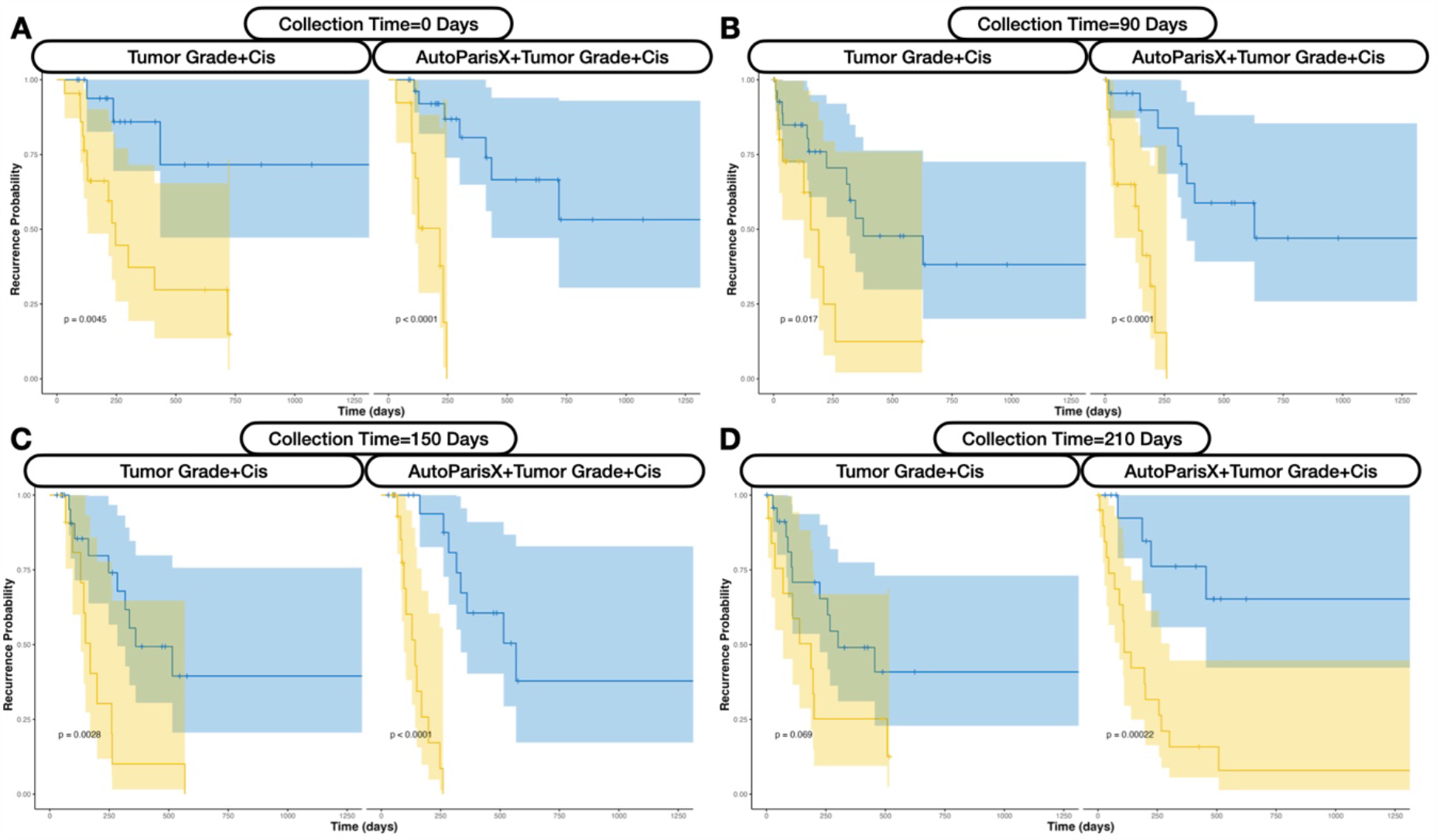
**Comparison of KM plots for Imaging versus Histological Predictors**, for cytological information collected across the following collection time periods after the first positive primary: **A)** 0 days, **B)** 90 days, **C)** 150 days, **D)** 210 days

**Supplementary Figure 3:**
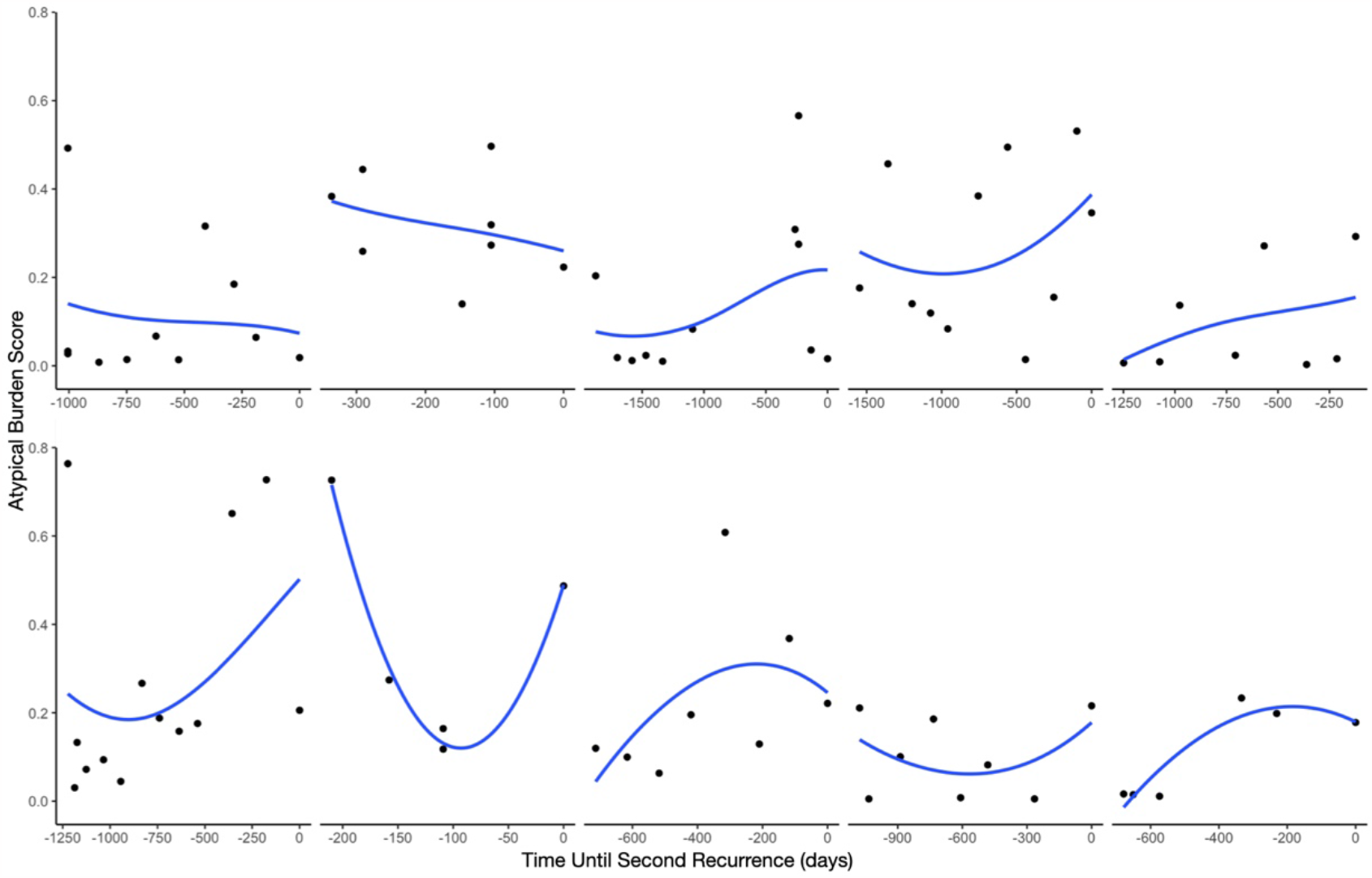
Atypia Burden Score Versus Time Until Second Recurrence: Reported for 10 patients with at least 4 repeat exams across the period between their first and second recurrence

**Supplementary Figure 4:**
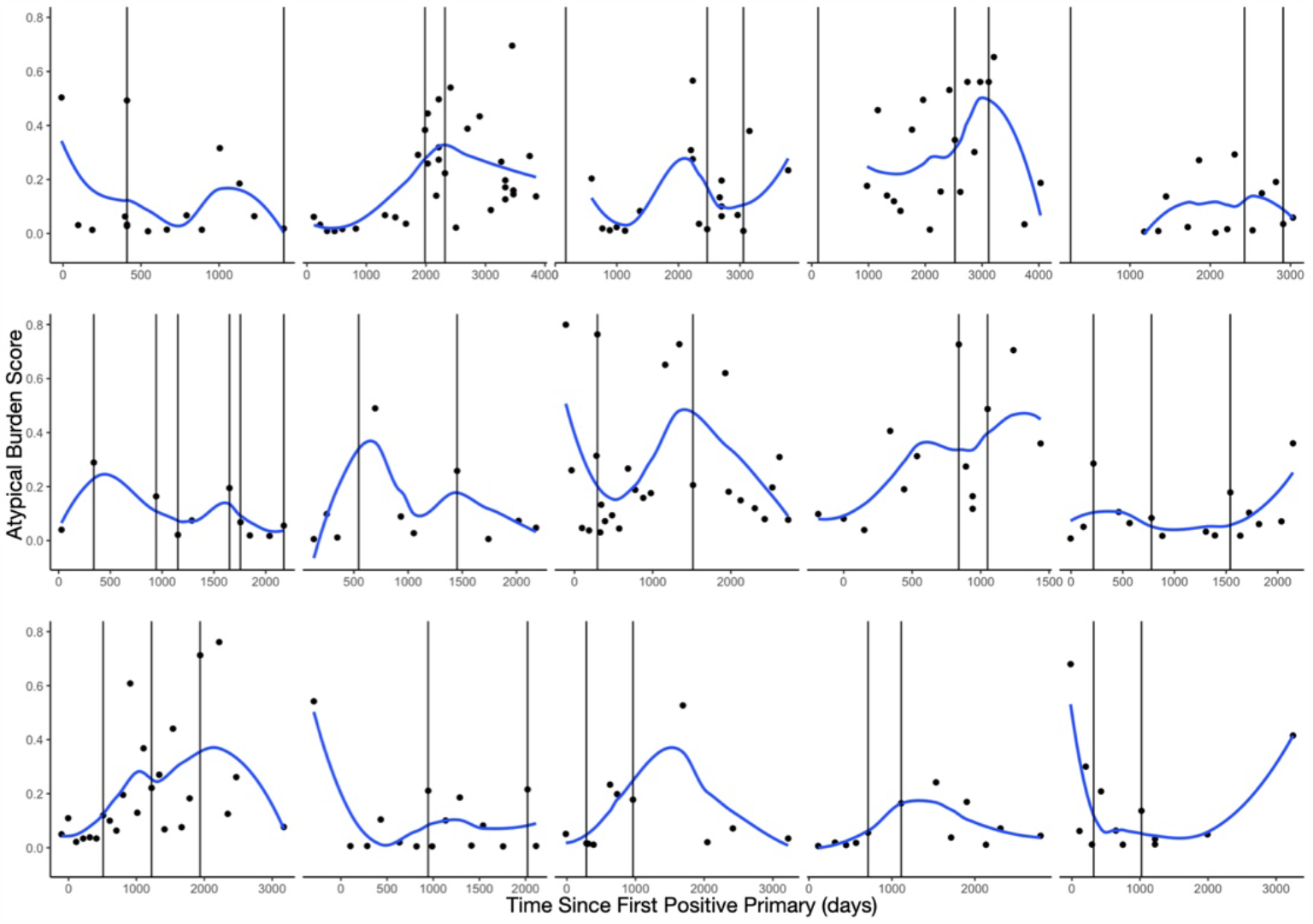
Atypia Burden Score Versus Time Across Multiple Recurrence Events for Select Patients: Each recurrence date is represented with the vertical line

**Supplementary Table 1:** Description of Slide Level predictors of Recurrence.

| Level | Predictor | Algorithm | Description |
| --- | --- | --- | --- |
| Cell | Urothelial cell score | UroNet | Predicted probability of urothelial cell from convolutional neural network, used to dynamically isolate urothelial cells in specimen |
|  | Atypia score | AtyNet | Predicted probability of presence of atypical features in urothelial cell (e.g., hyperchromasia, irregular nuclear membrane, etc.), determined using convolutional neural network |
|  | NC Ratio | UroSeg | Nuclear to cytoplasm area ratio derived from pixelwise segmentation of nucleus and cytoplasm using segmentation neural network |
|  | Morphometric measures | Custom | Complements binning of urothelial cells and assignment of atypia score, features: 1) area; 2) convex area; 3) eccentricity; 4) equivalent diameter; 5) extent; 6) Feret's diameter; 7) maximum diameter; 8) filled area; 9) major axis length; 10) minor axis length; 11) perimeter; and 12) solidity |
| Cluster | Dense Area | BorderDet | Whether cluster contains dense architecture of overlapping and indistinguishable cytoplasmic borders |
|  | Number urothelial cells | BorderDet/UroNet | Whether cluster contained urothelial cells, determined by counting cells with high urothelial cell score |
|  | Number atypical urothelial cells (atypia score) | BorderDet/UroNet/AtyNet | Whether cluster contained abnormal urothelial cells, determined by counting cells with high atypia score |
|  | Number atypical urothelial cells (NC ratio) | BorderDet/UroNet/UroSeg | Whether cluster contained abnormal urothelial cells, determined by counting cells with high NC ratio |
|  | Dense & Atypical | BorderDet/UroNet/AtyNet/UroSeg | Whether cluster contained both dense architecture and atypical cellular features |

| Slide | Patient characteristics | Supplied | Includes age, sex, history of hematuria, specimen source (e.g., voided), presence of specimen artifact |
| --- | --- | --- | --- |
|  | Isolated Cell-SIF Scores | Bayesian Optimization | Counting the number of cells with the following features from cells not associated with clusters: 1) cellularity (urothelial score), 2) atypia (atypia score), 3) atypia (NC ratio), 4) other morphometric measures |
|  | Cluster Cell-SIF Scores | Bayesian Optimization | Counting the number of cells with the following features from cells associated with clusters: 1) cellularity (urothelial score), 2) atypia (atypia score), 3) atypia (NC ratio), 4) other morphometric measures |
|  | All Cell-SIF Scores | Bayesian Optimization | Combines Isolated Cell-SIF Scores and Cluster Cell-SIF Scores |
|  | Cluster-SIF | Bayesian Optimization | Counting the number of clusters with the following features: 1) number of urothelial clusters, 2) atypical urothelial clusters (atypia score), 3) atypical clusters (NC ratio), 4) dense clusters, 5) dense and atypical clusters |
|  | Atypia Burden Score | Mixed effects machine learning | Integrates all slide-level predictors using machine learning model to calculate a score between 0-1 reflecting overall specimen atypia, correlated with UC diagnostic category |

**Supplementary Table 2:** Concordance statistics for *fixed predictors* at the following collection time periods; also included are performance statistics for *dynamic predictors* from the *time-varying covariate* cox model; the percentage of imaging variables which outperform manual examination is represented as “% Outperform UC Class”

| Collection Time (days) | 0 |  | 30 |  | 60 |  | 90 |  | 120 |  | 150 |  |
| --- | --- | --- | --- | --- | --- | --- | --- | --- | --- | --- | --- | --- |
| Predictors | C | SE | C | SE | C | SE | C | SE | C | SE | C | SE |
| ABS | 0.549 | 0.075 | 0.552 | 0.075 | 0.522 | 0.076 | 0.566 | 0.06 | 0.615 | 0.065 | 0.707 | 0.047 |
| # Dense Clusters | 0.62 | 0.069 | 0.62 | 0.069 | 0.6 | 0.069 | 0.581 | 0.061 | 0.666 | 0.066 | 0.74 | 0.051 |
| UC Class | 0.544 | 0.081 | 0.548 | 0.08 | 0.536 | 0.079 | 0.575 | 0.059 | 0.614 | 0.058 | 0.701 | 0.065 |
| Eccentricity | 0.558 | 0.084 | 0.564 | 0.088 | 0.515 | 0.078 | 0.557 | 0.059 | 0.662 | 0.059 | 0.716 | 0.048 |
| # Isolated Atypical Cells | 0.56 | 0.077 | 0.56 | 0.077 | 0.524 | 0.077 | 0.562 | 0.059 | 0.615 | 0.064 | 0.704 | 0.042 |
| # Atypical Clusters | 0.56 | 0.076 | 0.56 | 0.076 | 0.522 | 0.078 | 0.563 | 0.06 | 0.626 | 0.065 | 0.716 | 0.054 |
| # Overall Atypical Cells | 0.562 | 0.079 | 0.56 | 0.077 | 0.524 | 0.076 | 0.565 | 0.059 | 0.623 | 0.063 | 0.701 | 0.047 |
| # Cluster Atypical Cells | 0.558 | 0.076 | 0.556 | 0.075 | 0.522 | 0.076 | 0.561 | 0.059 | 0.623 | 0.062 | 0.698 | 0.047 |
| % Clusters Dense/Atypical | 0.554 | 0.076 | 0.554 | 0.076 | 0.519 | 0.077 | 0.564 | 0.06 | 0.631 | 0.051 | 0.683 | 0.054 |
| # Isolated Cells High NC | 0.558 | 0.081 | 0.558 | 0.081 | 0.522 | 0.076 | 0.564 | 0.058 | 0.617 | 0.063 | 0.713 | 0.047 |
| # Overall Cells High NC | 0.56 | 0.08 | 0.56 | 0.08 | 0.526 | 0.077 | 0.566 | 0.059 | 0.616 | 0.063 | 0.713 | 0.047 |
| # Cluster Cells High NC | 0.562 | 0.077 | 0.562 | 0.077 | 0.522 | 0.08 | 0.562 | 0.059 | 0.62 | 0.062 | 0.71 | 0.046 |
| LASSO | 0.59 | 0.084 | 0.603 | 0.073 | 0.578 | 0.069 | 0.584 | 0.06 | 0.654 | 0.058 | 0.74 | 0.051 |
| # Cells | 0.558 | 0.08 | 0.558 | 0.08 | 0.524 | 0.078 | 0.573 | 0.065 | 0.628 | 0.065 | 0.71 | 0.048 |
| # Clusters | 0.567 | 0.073 | 0.571 | 0.072 | 0.582 | 0.07 | 0.593 | 0.061 | 0.645 | 0.067 | 0.716 | 0.05 |
| Overall | 0.672 | 0.073 | 0.725 | 0.055 | 0.714 | 0.056 | 0.707 | 0.061 | 0.81 | 0.074 | 0.824 | 0.045 |
| VIF | 0.62 | 0.073 | 0.627 | 0.071 | 0.632 | 0.066 | 0.614 | 0.055 | 0.7 | 0.051 | 0.752 | 0.058 |
| % Outperform UC Class | 1.000 | 0.000 | 1.000 | 0.000 | 0.278 | 0.106 | 0.278 | 0.106 | 1.000 | 0.000 | 0.889 | 0.074 |

| Collection Time (days) | 180 |  | 210 |  | 240 |  | 270 |  | 300 |  | Time Varying Covariates |  |
| --- | --- | --- | --- | --- | --- | --- | --- | --- | --- | --- | --- | --- |
| Predictors | C | SE | C | SE | C | SE | C | SE | C | SE | C | SE |
| ABS | 0.722 | 0.048 | 0.708 | 0.051 | 0.689 | 0.063 | 0.68 | 0.067 | 0.703 | 0.06 | 0.652 | 0.039 |
| # Dense Clusters | 0.748 | 0.05 | 0.688 | 0.058 | 0.727 | 0.057 | 0.725 | 0.055 | 0.727 | 0.066 | 0.603 | 0.038 |
| UC Class | 0.724 | 0.062 | 0.673 | 0.063 | 0.689 | 0.06 | 0.682 | 0.059 | 0.689 | 0.059 | 0.579 | 0.05 |
| Eccentricity | 0.724 | 0.048 | 0.697 | 0.059 | 0.665 | 0.073 | 0.639 | 0.076 | 0.699 | 0.068 | 0.607 | 0.034 |
| # Isolated Atypical Cells | 0.717 | 0.043 | 0.692 | 0.056 | 0.702 | 0.065 | 0.697 | 0.066 | 0.709 | 0.061 | 0.612 | 0.039 |
| # Atypical Clusters | 0.733 | 0.053 | 0.716 | 0.062 | 0.7 | 0.067 | 0.702 | 0.064 | 0.712 | 0.06 | 0.62 | 0.039 |
| # Overall Atypical Cells | 0.715 | 0.048 | 0.679 | 0.062 | 0.685 | 0.069 | 0.682 | 0.068 | 0.663 | 0.067 | 0.638 | 0.04 |
| # Cluster Atypical Cells | 0.713 | 0.048 | 0.681 | 0.064 | 0.685 | 0.069 | 0.68 | 0.069 | 0.651 | 0.068 | 0.637 | 0.042 |
| % Clusters Dense/Atypical | 0.701 | 0.055 | 0.695 | 0.066 | 0.641 | 0.069 | 0.649 | 0.072 | 0.669 | 0.071 | 0.588 | 0.041 |
| # Isolated Cells High NC | 0.724 | 0.046 | 0.664 | 0.061 | 0.709 | 0.066 | 0.69 | 0.066 | 0.689 | 0.063 | 0.588 | 0.039 |
| # Overall Cells High NC | 0.727 | 0.047 | 0.655 | 0.054 | 0.707 | 0.066 | 0.685 | 0.065 | 0.709 | 0.062 | 0.564 | 0.042 |
| # Cluster Cells High NC | 0.724 | 0.047 | 0.645 | 0.057 | 0.697 | 0.068 | 0.68 | 0.066 | 0.699 | 0.068 | 0.541 | 0.044 |
| LASSO | 0.734 | 0.056 | 0.723 | 0.054 | 0.707 | 0.061 | 0.691 | 0.051 | 0.726 | 0.064 | 0.657 | 0.038 |
| # Cells | 0.724 | 0.045 | 0.705 | 0.052 | 0.707 | 0.058 | 0.692 | 0.063 | 0.712 | 0.059 | 0.631 | 0.039 |
| # Clusters | 0.727 | 0.049 | 0.702 | 0.053 | 0.743 | 0.051 | 0.719 | 0.056 | 0.677 | 0.067 | 0.6 | 0.039 |
| Overall | 0.827 | 0.046 | 0.849 | 0.051 | 0.927 | 0.035 | 0.92 | 0.036 | 0.911 | 0.03 | 0.659 | 0.041 |
| VIF | 0.773 | 0.058 | 0.747 | 0.057 | 0.731 | 0.061 | 0.743 | 0.074 | 0.746 | 0.06 | 0.682 | 0.036 |
| % Outperform UC Class | 0.611 | 0.115 | 0.722 | 0.106 | 0.667 | 0.111 | 0.611 | 0.115 | 0.667 | 0.111 | 0.778 | 0.098 |

**Supplementary Table 3:** Comparison between Cytological Imaging Predictors Versus Histology: Hazard ratios, 95% confidence intervals and p-values, specifically after adjusting for tumor grade/type, reported for a variable constructed from the imaging predictors alone; also includes p-values from partial likelihood ratio test assessing whether imaging cytological exams improves on histological predictors; reports for *fixed predictors* collected across various collection time periods

| Collection Time (days) | log(HR) | 2.5% CI | 97.5% CI | p-value | p-value– H1: Imaging> Grade+Cis | p-value– H1: Imaging+Grade+Cis> Grade+Cis |
| --- | --- | --- | --- | --- | --- | --- |
| 0 | 1.258 | 0.569 | 1.947 | 0.00035 | 0.220 | 0.048 |
| 30 | 1.199 | 0.533 | 1.865 | 0.00042 | 0.220 | 0.054 |
| 60 | 0.980 | 0.429 | 1.531 | 0.00049 | 0.060 | 0.042 |
| 90 | 0.982 | 0.319 | 1.646 | 0.00370 | 0.144 | 0.130 |
| 120 | 1.003 | 0.519 | 1.488 | 0.00005 | 0.115 | 0.102 |
| 150 | 1.019 | 0.538 | 1.499 | 0.00003 | 0.055 | 0.059 |
| 180 | 1.051 | 0.563 | 1.539 | 0.00002 | 0.060 | 0.060 |
| 210 | 1.081 | 0.542 | 1.621 | 0.00009 | 0.026 | 0.028 |
| 240 | 0.962 | 0.455 | 1.470 | 0.00020 | 0.024 | 0.017 |
| 270 | 0.974 | 0.517 | 1.432 | 0.00003 | 0.020 | 0.022 |
| 300 | 1.021 | 0.452 | 1.589 | 0.00044 | 0.016 | 0.017 |

**Supplementary Table 4:** C-indices for Imaging Predictors from *Time-Varying Effects*.

| Predictor | C | SE |
| --- | --- | --- |
| ABS | 0.65 | 0.039 |
| Age | 0.614 | 0.046 |
| # Dense Clusters | 0.578 | 0.037 |
| UC Class | 0.616 | 0.047 |
| Eccentricity | 0.563 | 0.049 |
| Sex | 0.54 | 0.041 |
| # Isolated Atypical Cells | 0.554 | 0.042 |
| # Atypical Clusters | 0.572 | 0.041 |
| # Overall Atypical Cells | 0.599 | 0.043 |
| % Clusters<br>Dense/Atypical | 0.568 | 0.043 |
| # Isolated Cells High NC | 0.558 | 0.039 |
| # Overall Cells High NC | 0.557 | 0.04 |
| # Cells | 0.603 | 0.044 |
| # Clusters | 0.627 | 0.042 |
| Overall | 0.728 | 0.043 |

**Supplementary Table 5:** Hazard Ratios for Imaging Predictors from Time Varying Effects Models; Predictor effect size and significance is reported for every half year, which was used as the time periods to assess recurrence risk

| Predictor | Time | log(HR) | 2.5% CI | 97.5% CI | z | Pr(>\|z\|) |
| --- | --- | --- | --- | --- | --- | --- |
| # Overall Atypical Cells | 0-180 | 1.39E-04 | 8.58E-05 | 1.93E-04 | 1.77E+00 | 7.63E-02 |
|  | 180-360 | 3.08E-04 | 1.90E-04 | 4.26E-04 | 1.51E+00 | 1.32E-01 |
|  | 360-540 | 4.60E-04 | 3.12E-04 | 6.07E-04 | 1.50E+00 | 1.33E-01 |
|  | 540-720 | 7.25E-04 | 1.53E-04 | 1.30E-03 | 6.43E-01 | 5.20E-01 |
|  | 720-900 | -6.14E-04 | -1.84E-03 | 6.08E-04 | -6.91E-01 | 4.89E-01 |
|  | >900 | 1.38E-03 | 8.03E-04 | 1.95E-03 | 1.10E+00 | 2.69E-01 |
| # Overall Cells High NC | 0-180 | 2.03E-04 | -2.87E-05 | 4.35E-04 | 6.93E-01 | 4.89E-01 |
|  | 180-360 | 8.75E-04 | 4.99E-04 | 1.25E-03 | 2.35E+00 | 1.89E-02 |
|  | 360-540 | 1.52E-03 | 9.92E-04 | 2.05E-03 | 1.29E+00 | 1.98E-01 |
|  | 540-720 | 1.09E-03 | -1.20E-04 | 2.30E-03 | 5.60E-01 | 5.75E-01 |
|  | 720-900 | -5.97E-05 | -2.13E-03 | 2.01E-03 | -3.43E-02 | 9.73E-01 |
|  | >900 | 8.15E-03 | 6.09E-03 | 1.02E-02 | 3.97E+00 | 7.23E-05 |
| # Cells | 0-180 | 4.72E-05 | 3.32E-05 | 6.11E-05 | 1.73E+00 | 8.39E-02 |
|  | 180-360 | 5.16E-05 | 3.65E-05 | 6.66E-05 | 1.65E+00 | 9.90E-02 |
|  | 360-540 | -4.94E-06 | -3.29E-05 | 2.31E-05 | -1.32E-01 | 8.95E-01 |
|  | 540-720 | 7.06E-05 | 4.95E-05 | 9.17E-05 | 2.49E+00 | 1.28E-02 |
|  | 720-900 | -1.07E-04 | -2.38E-04 | 2.33E-05 | -1.41E+00 | 1.58E-01 |
|  | >900 | 2.03E-04 | 1.43E-04 | 2.63E-04 | 1.58E+00 | 1.14E-01 |
| Eccentricity | 0-180 | 4.30E+00 | 1.89E+00 | 6.70E+00 | 8.73E-01 | 3.83E-01 |
|  | 180-360 | -1.60E+00 | -3.89E+00 | 6.87E-01 | -5.49E-01 | 5.83E-01 |
|  | 360-540 | 3.97E+00 | 4.59E-01 | 7.47E+00 | 4.50E-01 | 6.53E-01 |
|  | 540-720 | 1.72E+01 | 1.12E+01 | 2.31E+01 | 1.15E+00 | 2.49E-01 |
|  | 720-900 | 6.20E+00 | -2.94E+00 | 1.53E+01 | 3.73E-01 | 7.09E-01 |
|  | >900 | 5.72E+00 | -1.11E+00 | 1.25E+01 | 4.22E-01 | 6.73E-01 |
| # Isolated Atypical Cells | 0-180 | 1.21E-04 | -1.75E-04 | 4.17E-04 | 3.89E-01 | 6.98E-01 |
|  | 180-360 | 4.85E-04 | 1.04E-04 | 8.67E-04 | 8.19E-01 | 4.13E-01 |
|  | 360-540 | 2.02E-03 | 1.60E-03 | 2.44E-03 | 3.47E+00 | 5.22E-04 |
|  | 540-720 | 1.15E-04 | -2.23E-03 | 2.46E-03 | 2.37E-02 | 9.81E-01 |
|  | 720-900 | -4.80E-03 | -9.49E-03 | -1.21E-04 | -7.92E-01 | 4.29E-01 |
|  | >900 | 2.19E-03 | 1.24E-04 | 4.25E-03 | 5.51E-01 | 5.82E-01 |
| # Isolated Cells High NC | 0-180 | 3.20E-04 | -3.19E-04 | 9.60E-04 | 3.78E-01 | 7.05E-01 |
|  | 180-360 | 2.70E-03 | 1.86E-03 | 3.54E-03 | 3.79E+00 | 1.51E-04 |
|  | 360-540 | 2.33E-03 | 1.48E-03 | 3.18E-03 | 1.19E+00 | 2.34E-01 |
|  | 540-720 | 4.15E-03 | 2.00E-03 | 6.30E-03 | 1.24E+00 | 2.13E-01 |
|  | 720-900 | -2.49E-03 | -7.73E-03 | 2.75E-03 | -6.05E-01 | 5.45E-01 |
|  | >900 | 1.43E-02 | 1.09E-02 | 1.77E-02 | 2.95E+00 | 3.15E-03 |
| # Dense Clusters | 0-180 | 2.85E-04 | -5.81E-04 | 1.15E-03 | 2.38E-01 | 8.12E-01 |
|  | 180-360 | 2.08E-03 | 1.54E-03 | 2.61E-03 | 4.87E+00 | 1.12E-06 |
|  | 360-540 | 9.12E-03 | 7.59E-03 | 1.06E-02 | 4.20E+00 | 2.64E-05 |
|  | 540-720 | -1.02E-02 | -1.91E-02 | -1.20E-03 | -9.38E-01 | 3.48E-01 |
|  | 720-900 | -4.12E-03 | -1.39E-02 | 5.63E-03 | -3.27E-01 | 7.44E-01 |
|  | >900 | -3.12E-03 | -1.12E-02 | 4.96E-03 | -2.68E-01 | 7.89E-01 |
| # Clusters | 0-180 | 8.60E-05 | 5.55E-05 | 1.16E-04 | 1.56E+00 | 1.20E-01 |
|  | 180-360 | 1.39E-04 | 1.10E-04 | 1.69E-04 | 3.66E+00 | 2.50E-04 |
|  | 360-540 | 4.19E-05 | -9.12E-06 | 9.29E-05 | 4.55E-01 | 6.49E-01 |
|  | 540-720 | 4.93E-05 | -1.16E-05 | 1.10E-04 | 4.64E-01 | 6.43E-01 |
|  | 720-900 | -5.40E-04 | -8.79E-04 | -2.02E-04 | -1.85E+00 | 6.48E-02 |
|  | >900 | 5.93E-05 | -2.11E-05 | 1.40E-04 | 3.87E-01 | 6.99E-01 |
| % Clusters<br>Dense/Atypical | 0-180 | 8.25E+00 | 5.08E+00 | 1.14E+01 | 1.93E+00 | 5.30E-02 |
|  | 180-360 | 5.06E+00 | -2.06E+00 | 1.22E+01 | 3.65E-01 | 7.15E-01 |
|  | 360-540 | 2.56E+01 | 1.99E+01 | 3.14E+01 | 2.64E+00 | 8.39E-03 |
|  | 540-720 | -2.20E+01 | -4.32E+01 | -7.23E-01 | -6.85E-01 | 4.93E-01 |
|  | 720-900 | 3.74E+01 | 2.15E+01 | 5.34E+01 | 1.67E+00 | 9.44E-02 |
|  | >900 | 2.29E+01 | 7.72E+00 | 3.81E+01 | 1.02E+00 | 3.07E-01 |
| # Atypical<br>Clusters | 0-180 | 7.07E-04 | 2.25E-04 | 1.19E-03 | 1.39E+00 | 1.65E-01 |
|  | 180-360 | 2.89E-03 | 1.76E-03 | 4.02E-03 | 1.53E+00 | 1.26E-01 |
|  | 360-540 | 3.58E-03 | 2.71E-03 | 4.45E-03 | 2.12E+00 | 3.40E-02 |
|  | 540-720 | 3.22E-03 | -5.21E-04 | 6.96E-03 | 3.99E-01 | 6.90E-01 |
|  | 720-900 | -6.12E-03 | -1.40E-02 | 1.74E-03 | -5.87E-01 | 5.57E-01 |
|  | >900 | 4.81E-03 | 9.55E-04 | 8.67E-03 | 6.08E-01 | 5.43E-01 |
| ABS | 0-180 | 2.74E+00 | 2.17E+00 | 3.31E+00 | 3.00E+00 | 2.69E-03 |
|  | 180-360 | 2.22E+00 | 1.57E+00 | 2.86E+00 | 1.72E+00 | 8.51E-02 |
|  | 360-540 | -1.86E-01 | -1.22E+00 | 8.45E-01 | -1.30E-01 | 8.96E-01 |
|  | 540-720 | 8.28E-01 | -5.97E-01 | 2.25E+00 | 2.75E-01 | 7.84E-01 |
|  | 720-900 | 2.39E+00 | 6.42E-01 | 4.15E+00 | 9.43E-01 | 3.46E-01 |
|  | >900 | 7.96E+00 | 6.45E+00 | 9.47E+00 | 3.31E+00 | 9.41E-04 |
| UC Class | 0-180 | 1.61E+00 | 1.35E+00 | 1.88E+00 | 3.10E+00 | 1.96E-03 |
|  | 180-360 | 8.31E-01 | 5.37E-01 | 1.13E+00 | 1.38E+00 | 1.68E-01 |
|  | 360-540 | -2.07E-01 | -7.32E-01 | 3.19E-01 | -2.03E-01 | 8.39E-01 |
|  | 540-720 | 4.70E-01 | -7.19E-02 | 1.01E+00 | 4.38E-01 | 6.61E-01 |
|  | 720-900 | 1.29E+00 | 6.80E-01 | 1.91E+00 | 1.39E+00 | 1.64E-01 |
|  | >900 | 1.49E+00 | 1.00E+00 | 1.98E+00 | 1.55E+00 | 1.21E-01 |

**Supplementary Table 6:** Results from beta regression models comparing recurrence risk to ABS scores during distinct time periods; Coefficients **B** represents differences in ABS scores between low and high risk patients at specific time periods; the final coefficient represents how ABS scores are changing over time between the first and second recurrences

| Comparison | Time Period | B | 2.5% CI | 97.5% CI | p-value |
| --- | --- | --- | --- | --- | --- |
| High vs low risk, days since positive primary | 0-113 | -0.297 | -1.169 | 0.575 | 0.506 |
|  | 114-204 | 0.134 | -1.212 | 1.479 | 0.846 |
|  | 205-295 | -0.806 | -1.849 | 0.238 | 0.133 |
|  | 295-412 | -1.038 | -1.888 | -0.187 | 0.019 |
|  | 413-690 | -1.186 | -1.957 | -0.416 | 0.003 |
| High vs low risk, days until first recurrence | >752 | -0.070 | -0.645 | 0.505 | 0.812 |
|  | 752-391 | 0.122 | -0.438 | 0.683 | 0.669 |
|  | 390-227 | -0.496 | -1.086 | 0.095 | 0.102 |
|  | 226-114 | 0.093 | -0.459 | 0.645 | 0.742 |
|  | 113-0 | -0.595 | -1.193 | 0.003 | 0.053 |
| Days until second recurrence, starting from first recurrence | Time in days (continuous) | 0.001 | 0.000 | 0.001 | 0.018 |

